# Canada’s loss of measles elimination after 27 years: a mathematical modelling analysis of the five major provincial outbreak waves in 2025–2026

**DOI:** 10.64898/2026.10.30.26366944

**Authors:** Chapin S. Korosec, Lily McChesney-Knight, Edward W. Thommes, Monica G. Cojocaru

## Abstract

**Background:** Canada’s loss of measles elimination status in 2025 highlighted the vulnerability of populations with concentrated immunity gaps. Provincial incidence alone cannot readily distinguish relatively intense transmission within a small susceptible population from transmission involving a larger population. We quantified these differences across five provincial measles waves in 2025-2026.

**Methods:** We fitted a common susceptible–exposed–infectious–recovered (SEIR) model to weekly reported measles incidence for Ontario in 2025 and Alberta and Manitoba in 2025 and 2026. Mean latent and infectious periods were fixed at 10 and 8 days. We estimated the effective reproduction number (***R***_**eff**,**0**_), effective susceptible population (***S***_**eff**,**0**_), and profiled the initial infected number to evaluate province and wave-specific primary case model estimates. We calculate the initial exponential growth rate, doubling time, and peak full width at half maximum (FWHM) to characterize early epidemic growth and the duration of elevated incidence around each wave’s peak.

**Findings:** In 2025, Ontario’s effective reproduction number, ***R***_**eff**,**0**_, was 1.9, with a model-estimated effective susceptible population, ***S***_**eff**,**0**_, of 3,101 individuals, an initial doubling time of 2.4 weeks, and a peak FWHM of 12.3 weeks. Alberta had a similar ***R***_**eff**,**0**_ of 1.9, with ***S***_**eff**,**0**_ of 2,601 individuals, a doubling time of 2.4 weeks, and FWHM of 12.1 weeks. Manitoba had a lower ***R***_**eff**,**0**_ of 1.5 and ***S***_**eff**,**0**_ of 490 individuals, with a longer doubling time of 4.0 weeks and FWHM of 20.5 weeks. In 2026, Alberta’s ***R***_**eff**,**0**_ increased to 2.1 while ***S***_**eff**,**0**_ decreased to 383 individuals; doubling time shortened to 2.1 weeks and FWHM to 9.7 weeks. Manitoba’s ***R***_**eff**,**0**_ increased to 1.8 and ***S***_**eff**,**0**_ to 1,038 individuals, while doubling time shortened to 2.6 weeks and FWHM to 13.0 weeks.

**Interpretation:** Alberta’s smaller second wave combined the highest effective reproduction number with the shortest doubling time and peak duration, demonstrating that limited outbreak size can coexist with rapid transmission. Joint estimates of transmission intensity, epidemic pace, peak duration, and effective susceptible population provide complementary measures for comparing outbreaks and informing public-health planning. Together, these estimates provide a quantitative basis for distinguishing transmission intensity from the scale of susceptible involvement, supporting comparisons across outbreaks and informing the timing and scale of public-health responses for future outbreaks.

## Introduction

The 2025 global resurgence of measles included 14,891 confirmed cases in the Americas [1] and has been attributed to declining immunization coverage together with the resumption of pre-pandemic social mixing [2]. In Canada, the multijurisdictional outbreak resulted in 5,085 confirmed cases [3] with sustained transmission leading to The Pan American Health Organization to end the country’s 27-year measles elimination status [4]. This loss of elimination underscores a central challenge for measles control: national immunization averages can conceal geographically and socially concentrated pockets of susceptibility, making province-specific transmission models essential for quantifying regional epidemic dynamics [5]. Relating Canadian province-level estimates of transmission intensity, susceptible-population size, and outbreak seeding to the distribution of under-immunized subpopulations can identify where transmission was sustained, clarify how outbreaks became established across jurisdictions, and guide geographically targeted surveillance and immunization.

Measles is among the most transmissible human infections, with estimates of the reproduction number, *R*_0_, ranging from 12 to 18 and a corresponding theoretical herd-immunity threshold of approximately 92–94% in a homogeneously mixing population [6, 7]. However, vaccination decisions can cluster within social networks through shared beliefs, concentrating susceptible individuals within particular communities [8], while viral exposure is structured by age, household, school, and other social settings, with school-aged children exhibiting especially frequent and age-assortative contacts [9, 10]. Together, these processes connect susceptible individuals to one another more frequently than expected under random mixing, enabling localized outbreaks even when provincial or national vaccination coverage approaches the nominal herd-immunity threshold [8, 10]. Gromis and Liu [11] demonstrated through modelling of California schools that spatially clustered vaccine exemptions could sustain substantially larger measles outbreaks than randomly distributed exemptions at the same overall vaccination coverage, underscoring the importance of local immunity gaps. Although routine childhood vaccination remains widespread in Canada, population immunity is neither uniformly high nor evenly distributed. A national serosurvey using specimens collected from 2009 to 2013 found protective measles antibodies in 90.0% of Canadians, including only 78.7% of those aged 12–19 years, below levels generally considered necessary to sustain elimination [12]. More recently, Alberta data indicate that this vulnerability increased before the 2025 outbreak: first-dose MMR coverage by age two declined from 83.3% in 2021 to 80.1% in 2024, while fourth-dose DTaP-IPV-Hib coverage decreased from 72.5% to 68.9%, indicating that declining uptake extended beyond measles vaccination [13]. Further, in 2023, Alberta’s province-wide MMR coverage by age two was 81.7%, but estimates across 132 local geographic areas ranged from 24.6% to 91.1% [14]. These disparities can reflect vaccine hesitancy, delayed vaccination, and barriers to access, and are further obscured by Canada’s decentralized and incompletely harmonized immunization-surveillance systems [15, 16]. To address these gaps, Hadley et al. developed a Gaussian-process framework to estimate coverage with at least one measles vaccine dose across Canadian age groups and provinces or territories, providing inputs for outbreak modelling where surveillance data are incomplete [17]. A review of 80 Canadian measles outbreaks between 1999 and 2025 found that 79.7% of cases occurred among unvaccinated individuals and 61.7% of outbreaks were associated with imported infections, illustrating how repeated introductions can seed transmission within under-immunized communities [18]. Coverage data can therefore identify where immunization is low, but cannot determine the effective susceptible population exposed to, and participating in, transmission following viral introduction.

Mathematical transmission models bridge this gap by integrating reported incidence with measles natural history to infer otherwise unobserved epidemiological quantities, including transmission intensity, effective susceptibility, outbreak establishment, and the expected effects of interventions [19]. Recent modelling in highly vaccinated or near-elimination settings has primarily examined prospective scenarios or individual subnational outbreaks. In Canada, McNichol et al. [20] simulated introductions under varying vaccination coverage and intervention conditions, finding that when coverage exceeded 95%, 99% of introductions did not generate an outbreak. Ruma et al. extended measles transmission modelling to incorporate waning vaccine-induced immunity, examining its implications for vaccination thresholds and estimating the population involved in transmission during the 2024–2025 outbreak in Ontario’s Southwestern Public Health Region [21]. Subnational models have further demonstrated how case notifications can reconstruct variation in measles susceptibility across age, space, and time [5], while age- and region-stratified modelling in England has quantified how vaccination timing and coverage shape outbreak risk in a highly vaccinated population [22]. A two-region model calibrated to the 2025 outbreak in northern British Columbia additionally found that interregional mobility could increase outbreak probability by as much as 50%, demonstrating how connectivity can alter the effectiveness of local interventions [23]. A comparative analysis of the major provincial trajectories comprising Canada’s 2024–2025 multijurisdictional out-break remains needed to distinguish whether their observed case burdens arose from differences in transmission intensity, effective susceptible-population size, or the extent to which transmission was already established when observation began. Using an age-structured model of the 2018–2019 New York City measles outbreak, Yang [24] jointly inferred initial susceptibility and transmission parameters, highlighting how delayed vaccination and changes in infectious contact contributed to outbreak dynamics and estimating the substantial burden averted by vaccination campaigns. The analysis implicated delayed vaccination among 1–4-year-olds in early spread and increased infectious contact in later transmission, and estimated that infections and hospitalizations could have been more than tenfold higher without the vaccination campaigns [24].

In this study, we fitted a common susceptible–exposed–infectious–recovered modelling framework to five measles outbreak waves; Ontario (2025), Alberta (2025 & 2026), and Manitoba (2025 & 2026). We estimated the initial effective reproduction number, effective susceptible population, and initial infectious seed size, fixing latent and infectious periods in the main analysis and examining biologically plausible alternatives in sensitivity analyses. We additionally calculated the initial exponential growth rate (*r*), doubling time (*T*_d_), and peak full width at half maximum (FWHM) to characterize the pace of early spread and duration of elevated incidence. Together, these quantities distinguish how rapidly an outbreak expands from the size of the susceptible population sustaining it in the Canadian context, providing a quantitative basis for assessing response urgency and anticipating the scale and duration of public-health demands.

## Methods

### SEIR transmission model

We used a deterministic susceptible–exposed–infectious–recovered (SEIR) model with one exposed compartment,

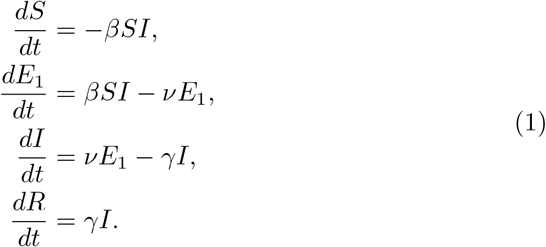

Time *t* was expressed in weeks, so that *β* has units of individuals^*−*1^ week^*−*1^ and *ν* and *γ* have units of week^*−*1^. The transmission coefficient was parameterized by the initial effective reproduction number,

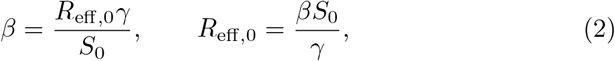

with initial conditions

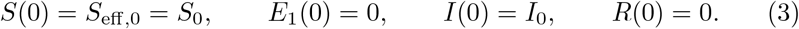

Here, *S*_eff,0_ represents the model-inferred effective susceptible population participating in the fitted outbreak, not the total number of susceptible provincial residents. The fitted *R*_eff,0_ is the initial effective reproduction number within this population, rather than the basic reproduction number in an entirely susceptible population. The quantity *I*_0_ represents infectious individuals present at the beginning of the fitted observation window and should not be interpreted as a count of independently introduced index cases.

### Incidence data

We assembled publicly available weekly measles incidence series for Ontario’s 2025 wave and Alberta’s and Manitoba’s 2025 and 2026 waves. Ontario reports were obtained from Public Health Ontario’s enhanced epidemiological summaries [25]; Alberta and Manitoba series were obtained from the Government of Canada’s 2025 and 2026 Measles and Rubella Weekly Monitoring Reports [26, 27]. Each wave was analysed independently, with *t* = 0 at the first observation in its respective fitted window and observation times *t*_*i*_ measured in weeks. Consistent with the implemented fitting specification, the model output compared with the reported weekly incidence series was the instantaneous incidence rate,

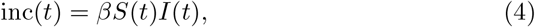

expressed in individuals week^*−*1^ and evaluated at the reported weekly observation times. This rate-based observation mapping is distinct from integrating incident infections over an entire reporting week.

### Parameter estimation and profile fitting

Model fitting was performed in Monolix [28] using the `lixoftConnectors` R library, with selected fits checked in the Monolix environment. For every candidate initial infectious seed size *I*_0_, and for each fixed pair of latent and infectious periods, the parameters *R*_eff,0_ and *S*_eff,0_ and additive and proportional residual-error parameters *a* and *b* were estimated by maximum likelihood. Standard errors and likelihood-based information criteria were obtained from the resulting fits. The five waves were fitted separately. Estimating an effective population participating in transmission has precedent in Ebola modelling, where Getz et al. [29] inferred the effective population at risk from weekly incidence rather than equating it with the full administrative population.

The reference analysis fixed the mean latent period at 10 days and the mean infectious period at 8 days. With model time in weeks, the corresponding transition rates were

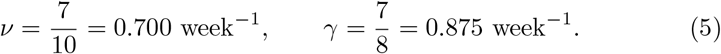

To investigate latent and infectious period model assumptions, supplementary sensitivity analyses considered all nine combinations

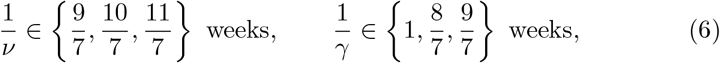

corresponding to latent periods of 9–11 days and infectious periods of 7–9 days. Initial seed size was examined using province-specific scans of candidate integer values, with additional fine-grid evaluations around minima. The audited reference-rate grids spanned *I*_0_ = 1–40 for Ontario, 1–402 for Alberta, and 1–102 for Manitoba. In the reference fits, neighboring integers around the supported minima were evaluated individually. For each wave and fixed (*ν, γ*) pair, profile support was defined using

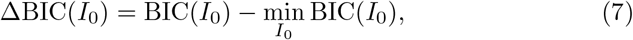

with evaluated fits satisfying ΔBIC *≤* 2 treated as comparably supported. The reported *I*_0_ support range consists of the smallest and largest supported evaluated seed sizes; it is not a confidence interval. The reference fit for each wave was the minimum-BIC fit for *ν* = 0.700 week^*−*1^ and *γ* = 0.875 week^*−*1^.

For the Ontario 2025 and Manitoba 2025 waves, *I*_0_ = 1 was also the lower boundary of the evaluated grid, an important qualification when interpreting their singleton support ranges.

### Uncertainty intervals for fitted trajectories

Best-fit trajectories were generated using each wave’s selected minimum-BIC parameter set. Standard errors for *R*_eff,0_ and *S*_0_ were obtained from the corresponding Monolix fit. Their relative standard errors (R.S.E.) were reported as

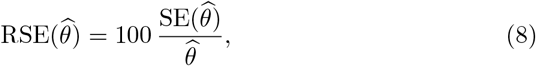

expressed as percentages and conditional on the fixed values of *ν, γ*, and *I*_0_. Approximate 95% Wald confidence intervals for *R*_eff,0_ and *S*_eff,0_ were calculated as 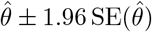.

Parameter uncertainty was propagated by Monte Carlo simulation around each wave’s selected minimum-BIC fit. For *m* = 1, …, 1000, we independently sampled

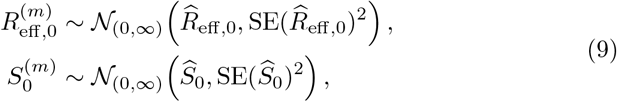

where *N*_(0,*∞*)_ denotes a normal distribution truncated at positive values, and the estimates and standard errors were obtained from the selected Monolix fit. For each draw, we recalculated 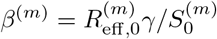 and simulated a trajectory with *ν, γ*, and *I*_0_ held fixed. This procedure generated an ensemble of simulated trajectories from a single selected fit per wave, rather than combining independently fitted models across the ΔBIC *≤* 2 support region. Point-wise 2.5%, 50%, and 97.5% quantiles summarized the resulting trajectories. The shaded bands in Figure 1A therefore represent approximate conditional parameter-uncertainty intervals. They exclude fitted-parameter covariance, residual observation error, and uncertainty in the fixed quantities, and are not full prediction intervals.

**Fig 1.**
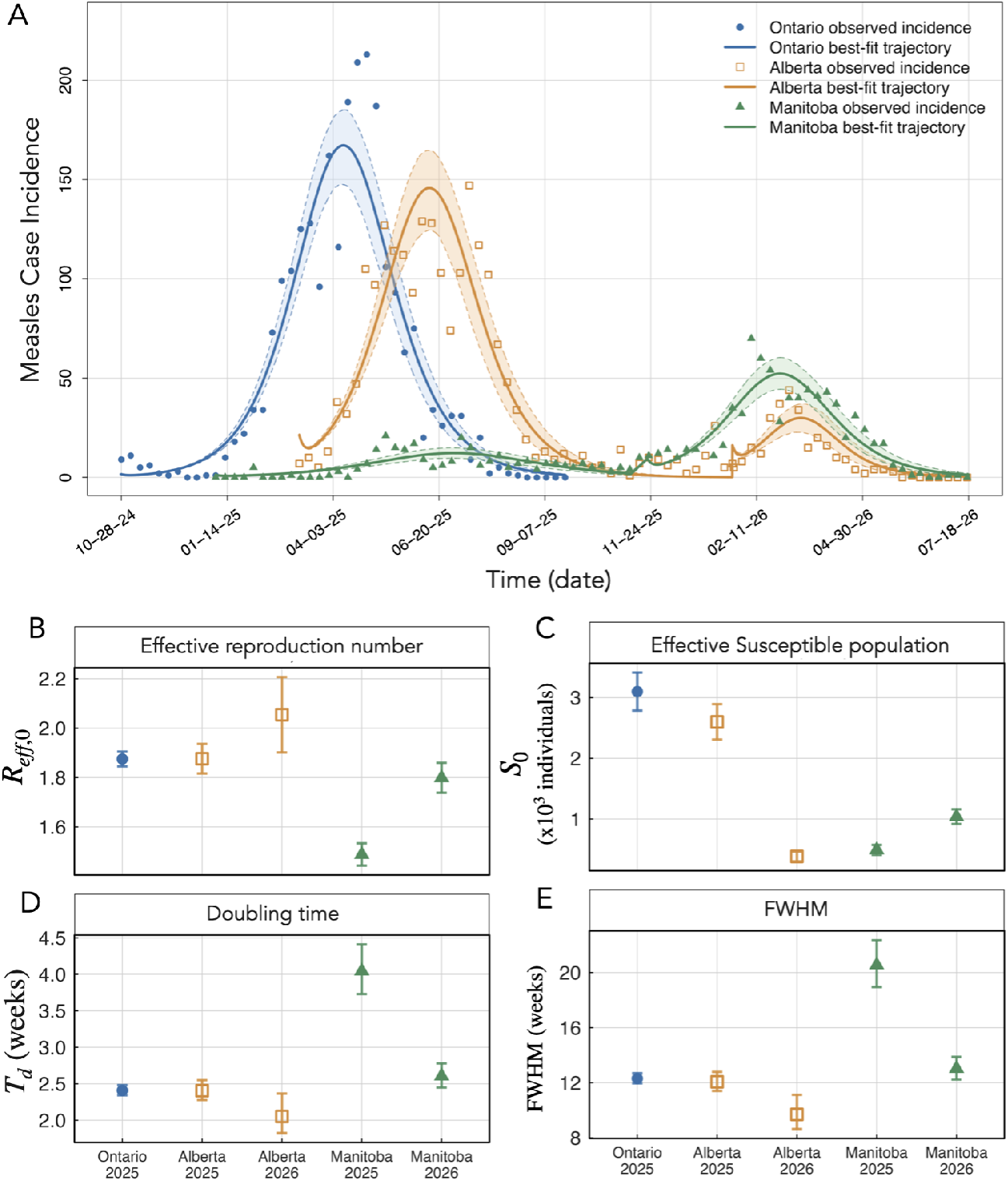
SEIR model fits and inferred outbreak quantities for the 2025–2026 measles waves in Ontario, Alberta, and Manitoba. A) Observed weekly measles case incidence and fitted trajectories for Ontario (2025), Alberta (2025 and 2026), and Manitoba (2025 and 2026). Shaded regions represent 95% conditional parameter-uncertainty bands propagated from estimated standard errors in *R*_eff,0_ and *S*_eff,0_. (B–E) Minimum-BIC model estimates of the effective reproduction number, susceptible population (thousands), initial doubling time (weeks), and peak full width at half maximum (weeks). Error bars show approximate 95% Wald confidence intervals in B–C and propagated 95% parameter-uncertainty intervals in D–E; FWHM intervals are conditional on draws with defined half-maximum crossings. Mean latent and infectious periods were fixed at 10 and 8 days, respectively.

### Derived growth and outbreak-duration measures

The initial effective reproduction number, *R*_eff,0_ = *βS*_0_*/γ*, is related to the dominant early exponential growth rate by

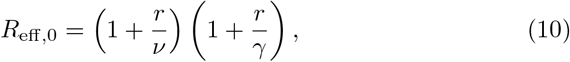

for the SEIR model with exponentially distributed latent and infectious periods [30]. We used this relationship to calculate the positive root

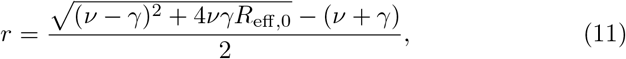

expressed in units of inverse weeks. The corresponding doubling time is therefore *T*_d_ = ln(2)*/r* weeks. We additionally measured the full width at half maximum (FWHM) of the fitted incidence trajectory as the interval, in weeks, between the rising- and falling-phase crossings of half the modelled peak incidence.

### Numerical implementation

Deterministic trajectories for plotting were solved using a fourth-order Runge– Kutta scheme. Expressed in weekly model units, the time steps described in the numerical implementation were

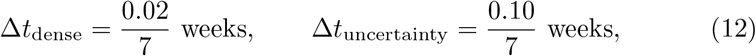

equivalent to 0.02 and 0.10 days, respectively. The Monolix fitting routine used population parameter estimation. A condensed version of the fit setup is provided in Table S1.

### Sensitivity analysis

For each wave, we performed Latin hypercube sampling with partial rank correlation coefficients (LHS–PRCC) using 2,000 parameter combinations per sampling design. Independent uniform ranges spanned *±*10% of fitted *R*_eff,0_ and *S*_0_, latent periods of 9–11 days, and infectious periods of 7–9 days. Initial infectious seed size *I*_0_ was fixed at its selected value, and *β* = *R*_eff,0_*γ/S*_0_ was recalculated for each simulation without refitting. PRCCs quantified associations with peak incidence, time to peak, FWHM, and initial doubling time after adjustment for the other sampled inputs. Approximate 95% percentile intervals were obtained from 500 paired bootstrap resamples. Three independent sampling designs and wider *±*20% ranges for *R*_eff,0_ and *S*_0_ assessed sampling and range sensitivity. Input–output plots supported checks of monotonicity.

## Results

The model fits described the broad rise, peak, and decline of five provincial measles waves (Fig. 1A). All fits described here used weekly incidence data and transition rates of *ν* = 0.700 week^*−*1^ and *γ* = 0.875 week^*−*1^, equivalent to mean latent and infectious periods of 10 and 8 days, with fit-sensitivity performed on these assumptions and results summarized in Table S2 and Figure S2. The fitted trajectories distinguished initial effective reproduction numbers, doubling time, and FWHM across all provincial measles waves (Fig. 1B–E; Table 1). Initial infected cases, *I*_0_, are profiled over a range of values for each fit to determine the minimal BIC *I*_0_ range for each provincial wave, the profile results and minimal BIC range are shown in Figure S1 and Table S2.

**Table 1.** Estimated transmission dynamics and outbreak characteristics for the Ontario 2025, Alberta 2025–2026, and Manitoba 2025–2026 measles waves. Reference fits fixed the mean latent and infectious periods at 10 and 8 days, respectively (*ν* = 0.700 week^*−*1^ and *γ* = 0.875 week^*−*1^). Estimates and relative standard errors (R.S.E.) correspond to each wave’s minimum-BIC fit. Initial infectious seed-size ranges span evaluated values satisfying ΔBIC *≤* 2 within each wave and represent model-profile support rather than confidence intervals. The singleton ranges for Ontario 2025 and Manitoba 2025 occur at the lower boundary of the evaluated seed-size grid. Growth rates, doubling times, and fitted-peak full widths at half maximum (FWHM) are reported in weekly units.

| **Quantity** | Ontario | Alberta |  | Manitoba |  |
|---|---|---|---|---|---|
| Year | 2025 | 2025 | 2026 | 2025 | 2026 |
| *R*<sub>eff,0</sub> (% R.S.E.) | 1.88 (0.81%) | 1.88 (1.63%) | 2.05 (3.79%) | 1.49 (1.55%) | 1.80 (1.72%) |
| *S*<sub>eff,0</sub> (% R.S.E.) | 3,101 (5.16%) | 2,601 (5.72%) | 383 (10.68%) | 490 (8.54%) | 1,038 (5.98%) |
| Initial infectious seed size, *I*<sub>0</sub> (support range) | 1 (1–1) | 13 (9–17) | 9 (6–12) | 1 (1–1) | 6 (5–7) |
| Initial exponential growth rate, *r* (week<sup>−1</sup>) | 0.288 | 0.288 | 0.338 | 0.171 | 0.266 |
| Initial doubling time estimate (weeks) | 2.41 | 2.41 | 2.05 | 4.05 | 2.61 |
| Peak FWHM (weeks) | 12.31 | 12.07 | 9.70 | 20.54 | 13.04 |
| Minimum BIC | 364 | 339 | 141 | 235 | 224 |

All five fits to Canadian provincial measles outbreaks in Ontario, Alberta and Manitoba yielded *R*_eff,0_ *>* 1 (Figure 1B, Table 1). In 2025, the fitted initial effective reproduction numbers were nearly identical in Ontario (1.88) and Alberta (1.88), compared with 1.49 for Manitoba. Their respective effective susceptible populations were 3,101, 2,601, and 490 individuals. For Alberta, *R*_eff,0_ increased from 1.88 in 2025 to 2.05 in 2026, while *S*_eff,0_ fell from 2,601 to 383 individuals, an approximately 85.3% reduction. Hence, the smaller estimated participating susceptible population in 2026 coincided with the highest initial effective reproduction number among the five waves. In Manitoba, *R*_eff,0_ increased from 1.49 in 2025 to 1.80 in 2026, while *S*_eff,0_ rose from 490 to 1,038 individuals, approximately 2.12 times its 2025 value.

The derived early exponential growth rate *r* ranged from 0.171 to 0.338 week^*−*1^ across the five waves (Table 1, with doubling times plotted in Figure 1D). Ontario’s and Alberta’s 2025 growth rates were similar (0.288 week^*−*1^ in each), with corresponding doubling times of approximately 2.41 weeks; Manitoba’s 2025 growth rate was 0.171 week^*−*1^, giving the longest doubling time (4.05 weeks). Alberta’s 2026 wave had the fastest estimated early growth (0.338 week^*−*1^), the shortest doubling time (2.05 weeks), and the narrowest fitted incidence peak (FWHM 9.70 weeks, compared with 12.07 weeks in 2025). Manitoba’s estimated growth rate rose to 0.266 week^*−*1^ in 2026, with doubling time decreasing to 2.61 weeks and fitted FWHM decreasing from 20.54 weeks in 2025 to 13.04 weeks in 2026. Ontario’s fitted 2025 FWHM was 12.31 weeks. These FWHM values summarize the durations of fitted incidence peaks rather than the complete durations of the outbreaks.

The minimum-BIC initial infectious seed sizes were *I*_0_ = 1 for Ontario 2025, *I*_0_ = 13 for Alberta 2025, *I*_0_ = 9 for Alberta 2026, *I*_0_ = 1 for Manitoba 2025, and *I*_0_ = 6 for Manitoba 2026 (Fig. S1 Table 1). Corresponding ΔBIC *≤* 2 support ranges were 1–1, 9–17, 6–12, 1–1, and 5–7. The singleton ranges for Ontario 2025 and Manitoba 2025 arise at the minimum evaluated seed size (*I*_0_ = 1) and therefore should not be interpreted as narrowly identified confidence intervals.

Across the BIC-minimizing fits, relative standard errors ranged from 0.81% to 3.79% for *R*_eff,0_ and from 5.16% to 10.68% for *S*_eff,0_. Alberta’s 2026 fit had the largest relative standard error for both quantities. The minimum BICs for Ontario 2025, Alberta 2025, Alberta 2026, Manitoba 2025, and Manitoba 2026 were 364, 339, 141, 235, and 224, respectively.

Across all five waves, LHS–PRCC analysis associated higher *R*_eff,0_ with; greater peak incidence, earlier peaks, shorter FWHM, and shorter doubling times (Fig. S3). Larger *S*_0_ was strongly associated with greater peak incidence but showed much weaker associations with FWHM and negligible associations with doubling time. Outbreak wave-specific input–output diagnostics are shown in Figs. S4–S8.

## Discussion

National estimates of two-dose measles vaccine coverage by age seven decreased from 85.7% in 2013 to 79.2% in 2021 [15]. Together with the theoretical 92–94% herd-immunity threshold under homogeneous mixing [6, 7], this background underscores the potential importance of geographically and socially concentrated measles susceptibility within Canada. Using a common SEIR framework fitted to weekly observations, we characterized the initial effective reproduction number, effective susceptible-population size, and initial infectious seed size across five major provincial measles waves during Canada’s 2025–2026 resurgence. In all waves, the fitted *R*_eff,0_ exceeded unity, ranging from 1.49 (Manitoba 2025) to 2.05 (Alberta 2026). The 2025 Ontario and Alberta waves had identical *R*_eff,0_ estimates (1.88), as well as initial exponential growth rates and peak FWHM, despite different fitted effective susceptible populations.

The effective susceptible populations represent the model-estimated size of the populations participating in the outbreaks. For 2025; Ontario and Alberta had estimates of approximately 3,101 and 2,601 individuals, respectively, compared with 490 for Manitoba. Differences across waves may reflect changes in the connected populations reached by transmission as well as changes in contact patterns, case reporting, or interventions. The two Alberta measles waves provide the clearest contrast between transmission intensity and participating population size. The estimated *S*_eff,0_ declined from 2,601 in 2025 to 383 in 2026, a reduction of about 85%, while *R*_eff,0_ increased from 1.88 to 2.05. Alberta’s 2026 wave also had the fastest estimated early exponential growth (0.338 week^*−*1^), shortest doubling time (2.05 weeks), and narrowest incidence peak (FWHM 9.70 weeks). Thus, a wave involving a smaller fitted effective susceptible population need not indicate less intense early transmission. By contrast, Manitoba experienced increases in model epidemiological parameters from 2025 to 2026: *S*_eff,0_ rose from 490 to 1,038 (approximately 2.12-fold), and *R*_eff,0_ rose from 1.49 to 1.80. Conversely, Manitoba measles outbreak estimated doubling time fell from 4.05 to 2.61 weeks and fitted FWHM from 20.54 to 13.04 weeks (Table 1). Initial growth rates translate transmission potential into the timescale over which an outbreak exponentially expands: the estimated doubling times of 2.05–4.05 weeks indicate how quickly incidence could increase during early exponential growth. The importance of timely intervention is illustrated by the 2014 measles outbreak in an Amish community in Ohio, where cases were estimated to double approximately every five days during early spread, while the effective reproduction number declined from approximately 4 to 1 within two weeks as containment measures began [31]. Although our estimated doubling times were longer (2.05–4.05 weeks), both analyses highlight the importance of initiating control measures during early growth, before successive generations of transmission increase the burden of response. Our estimates provide a practical timescale for surveillance and response planning, while FWHM characterizes the duration of elevated incidence around the fitted peak. These results are consistent with differences in the size and connectivity of populations reached by transmission, however, do not establish whether depletion of susceptibility, vaccination hesitancy, repeat introduction, changing contacts, altered reporting, or outbreak control measures caused the changes.

Historical Canadian elimination-era estimates provide context to the recent outbreak but require careful comparison. King et al. [32] reported national effective reproduction numbers based on imported-case proportions of 0.58, 0.72, 0.95, and 0.62 for outbreaks occurring in 1998–2001, following the adoption of two-dose vaccination schedules and catch-up campaigns in 1996– 1997. King et al. [32] describe transmission in under-immunized religious communities even during elimination [32]. More recent descriptions of the 2025–2026 resurgence similarly emphasize transmission among closely connected under-vaccinated communities [16]. Using early outbreak data, Moore and Fisman [33] estimated reproduction numbers of approximately 1.51 nationally and 1.67 for Ontario within the modelled misinformation-associated subgroup under selected formulations, broadly comparable in magnitude to our *R*_*eff*,0_ estimates (see Table 1) but based on a different population structure and fitting approach. Such observations motivate investigating local immunity gaps and, together with our findings, support targeted public-health efforts to improve vaccine uptake within under-immunized communities and interrupt sustained transmission within small connected communities.

Our model-determined initial infectious seed sizes in 2025 clarify that a small number of individuals can seed a provincial outbreak. The 2025 estimates for *I*_0_ is one for Ontario, thirteen for Alberta, and one in Manitoba. For Ontario, this result is supported by Ontario Public Health’s announcement that the outbreak in the province was seeded by the New Brunswick cluster, which began from an international travel-related importation from one infectious individual. Our model results support that a single individual may be responsible for travel from New Brunswick to seed the outbreak in Ontario. These findings underscore the value of surveillance that detects small transmission clusters, timely information sharing across jurisdictions, and rapid case investigation, contact tracing, and targeted vaccination to limit subsequent spread.

We caution that model estimates in this work are conditional on a homogeneous-mixing SEIR model with constant parameters and the reported incidence data. Changes in ascertainment, contact patterns, and interventions may therefore be absorbed into the fitted parameters, and *S*_eff,0_ should not be interpreted as province-wide susceptibility, but rather as the model-estimated number of susceptible individuals participating in transmission within the fitted outbreak wave.

Models incorporating waning immunity and immune boosting have demonstrated that recurrent measles outbreaks can arise even under high vaccination uptake, highlighting how changes in population immunity can shape epidemic dynamics beyond the single-wave framework considered here [34]. Linking wave-specific transmission estimates with geographically resolved vaccination coverage, contact patterns, and population mobility could help identify where susceptibility persists and how communities remain connected by transmission. Such integration could support timely outbreak response by informing ongoing immunization efforts and identifying where additional surveillance and vaccination outreach are needed while the outbreak is occurring. Realizing this potential will require collaboration with public-health agencies to provide timely, geographically resolved surveillance and vaccination data to calibrate and validate models as outbreaks unfold.

## Data Availability

Data used in this manuscript is available github [35]

## Code Availability

Code used in this manuscript is available github [35]

## Acknowledgements

M.G.C acknowledges funding in support of this work from the INSPIRE CBRF grant 431005.

## Author Contributions

Conceptualization: CSK, Data Curation: CSK, LMK, Formal Analysis: CSK, LMK, Funding Acquisition: CSK, MGC, Methodology: CSK, LMK, EWT, MGC, Project Administration: CSK, MGC, Supervision: CSK, MGC, Validation: CSK, EWT, Visualization: CSK, Writing – Original Draft: CSK, Writing – Review & Editing: All authors

## Additional Information

## Correspondance

The requests for materials should be addressed to C.S.K.

