## Supplementary material for "Canada’s loss of measles elimination after 27 years: a mathematical modelling analysis of the five major provincial outbreak waves in 2025–2026"

### Contents

**1 Condensed summary of numerical implementation 2**

**2 Profiling $I_0$ to determine a minimal BIC range for each provincial wave 3**

**3 Sensitivity of model fits to fixed values of $\nu$ and $\gamma$ 4**

**4 Latin Hypercube Sampling (LHS) paired with Partial Rank Correlation Coefficient (PRCC) 6**

**5 LHS-PRCC input/output diagnostics 7**

**6 Correspondance 12**

### 1 Condensed summary of numerical implementation

| Quantity | Definition | Status | Units / values |
|---|---|---|---|
| $S_0 = S_{\mathrm{eff},0}$ | Initial effective susceptible population | Fitted | Individuals |
| $E_1(0)$ | Initial exposed population | Fixed | 0 individuals |
| $I_0$ | Initial infectious seed size | Profiled; fixed within each fit | Individuals |
| $R(0)$ | Initial recovered population | Fixed | 0 individuals |
| $R_{\mathrm{eff},0}$ | Initial effective reproduction number | Fitted | Dimensionless |
| $\beta$ | Transmission coefficient | Derived | $R_{\mathrm{eff},0}\gamma/S_0$; individuals$^{-1}$ week$^{-1}$ |
| $\nu$ | Exposed-to-infectious transition rate | Fixed within each fit | 0.700 week$^{-1}$ in the main analysis |
| $L = 1/\nu$ | Mean latent period | Fixed; varied in sensitivity analysis | Main: 10/7 weeks; sensitivity: $9/7, 10/7, 11/7$ weeks |
| $\gamma$ | Infectious-to-recovered transition rate | Fixed within each fit | 0.875 week$^{-1}$ in the main analysis |
| $D = 1/\gamma$ | Mean infectious period | Fixed; varied in sensitivity analysis | Main: 8/7 weeks; sensitivity: $1, 8/7, 9/7$ weeks |
| $a$ | Additive residual error parameter | Fitted | Same units as incidence observations |
| $b$ | Proportional residual error parameter | Fitted | Dimensionless |
| $\mathrm{inc}(t)$ | Model incidence output | Matched to observed incidence | $\beta S(t)I(t)$; individuals week$^{-1}$ |
| BIC | Bayesian information criterion | Calculated for each fitted model | Minimum identifies the selected fit |
| $\Delta$BIC | Difference from the minimum BIC | Derived within each wave and $(L, D)$ combination | $\mathrm{BIC} - \mathrm{BIC}_{\mathrm{min}}$ |
| $I_0$ support range | Minimum and maximum supported seed sizes | Derived from evaluated profile fits | Evaluated $I_0$ values with $\Delta\mathrm{BIC} \leq 2$ |
| R.S.E. | Relative standard error of a fitted parameter $\theta$ | Estimated for the selected fit | $100\,\mathrm{SE}(\hat{\theta})/\hat{\theta}$; % |
| $r$ | Initial exponential growth rate | Derived from the selected fit | $\dfrac{\sqrt{(\nu-\gamma)^2 + 4\nu\gamma R_{\mathrm{eff},0}} - (\nu+\gamma)}{2}$; week$^{-1}$ |
| $T_{\mathrm{d}}$ | Initial doubling time | Derived | $\ln(2)/r$; weeks, for $r > 0$ |
| FWHM | Duration between crossings of half the peak incidence | Derived from the selected fitted trajectory | Weeks |

**Table S1** SEIR model quantities and estimation procedures for the Ontario 2025, Alberta 2025–2026, and Manitoba 2025–2026 measles waves. Each wave was fitted independently using a single exposed compartment and weekly incidence observations. All model rates are expressed in inverse weeks. The main analysis fixed the exposed-to-infectious and infectious-to-recovered transition rates at $\nu = 0.700$ week$^{-1}$ and $\gamma = 0.875$ week$^{-1}$, corresponding to mean latent and infectious periods of 10/7 and 8/7 weeks, respectively. Initial infectious seed size was profiled, with $S_{\mathrm{eff},0}$, $R_{\mathrm{eff},0}$, and residual error parameters re-estimated at each evaluated $I_0$. Supplementary sensitivity analyses considered all nine combinations of $L \in \{9/7, 10/7, 11/7\}$ and $D \in \{1, 8/7, 9/7\}$ weeks. BIC minima and support ranges were determined separately for each wave and period combination. Profile support ranges were defined using $\Delta\mathrm{BIC} \leq 2$ and are not confidence intervals. The growth rate describes the early exponential regime before appreciable susceptible depletion, while FWHM summarizes the duration of the fitted incidence peak above half its maximum value.

### 2 Profiling $I_0$ to determine a minimal BIC range for each provincial wave

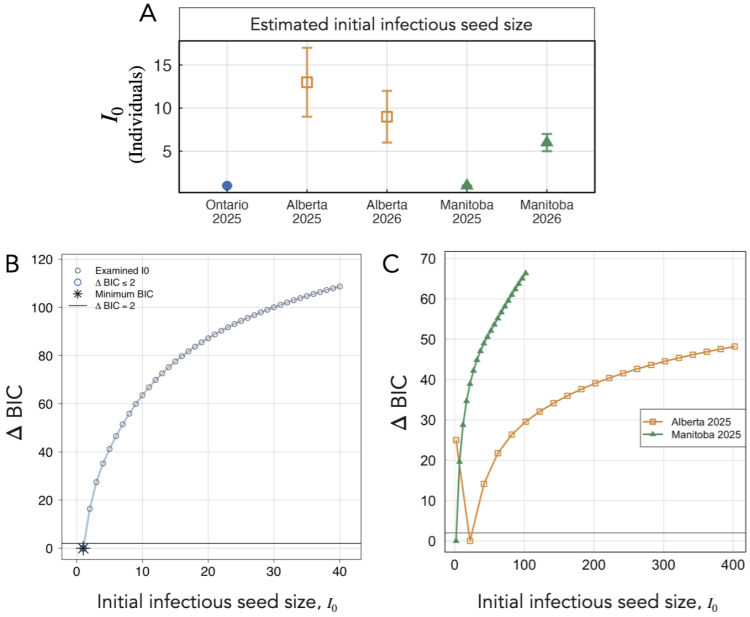


**Fig. S1 Initial infectious seed-size estimates and model support.** (A) Minimum-BIC estimates of $I_0$ for Ontario 2025, Alberta 2025–2026, and Manitoba 2025–2026. Error bars span evaluated seed sizes satisfying $\Delta\text{BIC} \leq 2$ and represent model-support ranges, not confidence intervals. (B) Ontario 2025 and (C) Alberta and Manitoba 2025 seed-size profiles; horizontal lines indicate $\Delta\text{BIC} = 2$, and the asterisk in B marks the minimum. Mean latent and infectious periods were fixed at 10 and 8 days, respectively, with other fitted parameters re-estimated at each $I_0$. The singleton support ranges for Ontario 2025 and Manitoba 2025 occur at the lowest evaluated seed size, $I_0 = 1$.

### 3 Sensitivity of model fits to fixed values of $\nu$ and $\gamma$

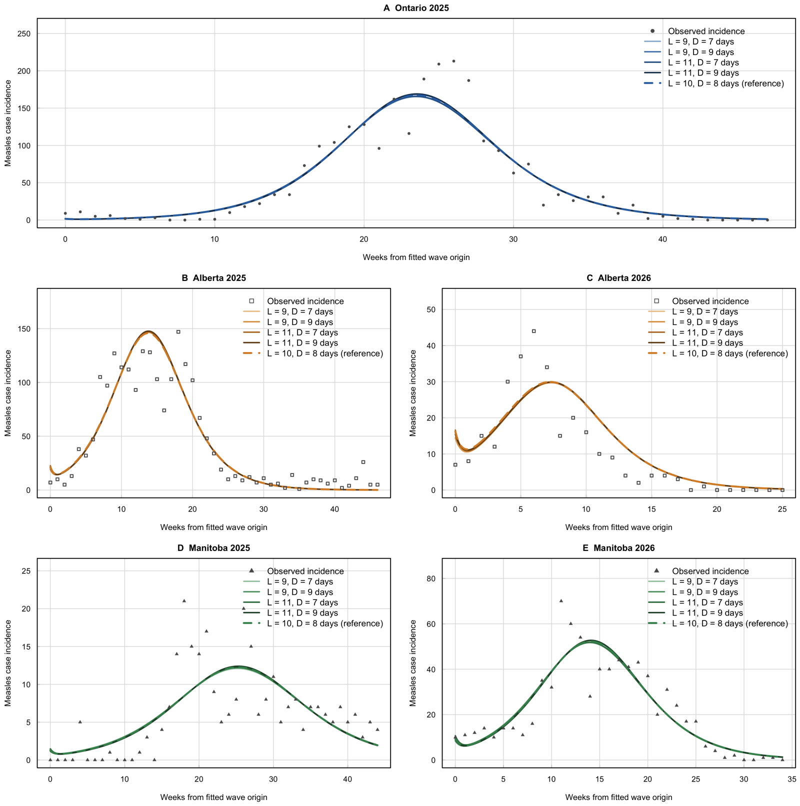


**Fig. S2 Sensitivity of fitted measles trajectories to latent and infectious periods.** Observed incidence and fitted SEIR trajectories for Ontario 2025 (A), Alberta 2025 (B), Alberta 2026 (C), Manitoba 2025 (D), and Manitoba 2026 (E). Grey symbols denote observations; solid curves show fits for mean latent and infectious periods $(L, D)$ of $(9, 7)$, $(9, 9)$, $(11, 7)$, and $(11, 9)$ days, respectively, shaded from light to dark. Dashed curves indicate the reference values $(L, D) = (10, 8)$ days. Each trajectory uses the minimum-BIC initial infectious seed size, $I_0$, and corresponding re-estimated parameters within its period combination. The closely overlapping curves (indistinguishable from the dashed reference) indicate that fitted peak timing, magnitude, and overall wave shape are relatively insensitive to these period choices after refitting; this agreement does not imply that the underlying parameter estimates are unchanged. Time is measured from each fitted wave's origin, and axis ranges vary between panels.

| Province | Year | Latent period (days) | Infectious period (days) | Minimum BIC | $I_0$ at minimum BIC | Accepted $I_0$ range | $S_0$ at minimum BIC | $S_0$ RSE (%) | $R_{\text{eff},0}$ at minimum BIC | $R_{\text{eff},0}$ RSE (%) |
|---|---|---|---|---|---|---|---|---|---|---|
| Ontario | 2025 | 9 | 7 | 365 | 1 | 1–1 | 3284 | 5.2 | 1.76 | 0.76 |
| Ontario | 2025 | 9 | 8 | 364 | 1 | 1–1 | 3182 | 5.2 | 1.82 | 0.79 |
| Ontario | 2025 | 9 | 9 | 363 | 1 | 1–1 | 3096 | 5.1 | 1.88 | 0.81 |
| Ontario | 2025 | 10 | 7 | 365 | 1 | 1–1 | 3190 | 5.2 | 1.81 | 0.79 |
| Ontario | 2025 | 10 | 8 | 364 | 1 | 1–1 | 3101 | 5.2 | 1.88 | 0.81 |
| Ontario | 2025 | 10 | 9 | 363 | 1 | 1–1 | 3024 | 5.1 | 1.94 | 0.83 |
| Ontario | 2025 | 11 | 7 | 365 | 1 | 1–1 | 3110 | 5.2 | 1.87 | 0.82 |
| Ontario | 2025 | 11 | 8 | 364 | 1 | 1–1 | 3030 | 5.2 | 1.93 | 0.84 |
| Ontario | 2025 | 11 | 9 | 363 | 1 | 1–1 | 2962 | 5.1 | 2.00 | 0.85 |
| Alberta | 2025 | 9 | 7 | 339 | 12 | 9–16 | 2736 | 5.7 | 1.77 | 1.51 |
| Alberta | 2025 | 9 | 8 | 339 | 13 | 9–17 | 2670 | 5.7 | 1.82 | 1.58 |
| Alberta | 2025 | 9 | 9 | 339 | 13 | 10–18 | 2591 | 5.7 | 1.89 | 1.62 |
| Alberta | 2025 | 10 | 7 | 339 | 12 | 9–16 | 2659 | 5.8 | 1.83 | 1.57 |
| Alberta | 2025 | 10 | 8 | 339 | 13 | 9–17 | 2601 | 5.7 | 1.88 | 1.63 |
| Alberta | 2025 | 10 | 9 | 339 | 13 | 10–18 | 2532 | 5.8 | 1.95 | 1.69 |
| Alberta | 2025 | 11 | 7 | 340 | 12 | 9–16 | 2594 | 5.8 | 1.88 | 1.62 |
| Alberta | 2025 | 11 | 8 | 339 | 13 | 9–17 | 2542 | 5.8 | 1.93 | 1.66 |
| Alberta | 2025 | 11 | 9 | 339 | 13 | 10–18 | 2480 | 5.9 | 2.00 | 1.73 |
| Alberta | 2026 | 9 | 7 | 141 | 8 | 6–12 | 392 | 11.0 | 1.93 | 3.58 |
| Alberta | 2026 | 9 | 8 | 141 | 9 | 6–12 | 391 | 10.8 | 1.98 | 3.80 |
| Alberta | 2026 | 9 | 9 | 141 | 9 | 7–13 | 379 | 11.2 | 2.06 | 3.92 |
| Alberta | 2026 | 10 | 7 | 141 | 8 | 6–11 | 383 | 11.4 | 2.00 | 3.84 |
| Alberta | 2026 | 10 | 8 | 141 | 9 | 6–12 | 383 | 10.7 | 2.05 | 3.79 |
| Alberta | 2026 | 10 | 9 | 141 | 9 | 7–13 | 372 | 10.9 | 2.13 | 3.88 |
| Alberta | 2026 | 11 | 7 | 141 | 8 | 6–11 | 376 | 11.1 | 2.08 | 3.78 |
| Alberta | 2026 | 11 | 8 | 141 | 8 | 6–12 | 365 | 11.4 | 2.16 | 3.90 |
| Alberta | 2026 | 11 | 9 | 141 | 9 | 7–12 | 366 | 10.9 | 2.21 | 3.97 |
| Manitoba | 2025 | 9 | 7 | 235 | 1 | 1–1 | 537 | 8.4 | 1.42 | 1.42 |
| Manitoba | 2025 | 9 | 8 | 235 | 1 | 1–1 | 510 | 8.5 | 1.46 | 1.49 |
| Manitoba | 2025 | 9 | 9 | 235 | 1 | 1–1 | 488 | 8.6 | 1.49 | 1.55 |
| Manitoba | 2025 | 10 | 7 | 235 | 1 | 1–1 | 514 | 8.5 | 1.45 | 1.48 |
| Manitoba | 2025 | 10 | 8 | 235 | 1 | 1–1 | 490 | 8.5 | 1.49 | 1.55 |
| Manitoba | 2025 | 10 | 9 | 235 | 1 | 1–1 | 470 | 8.6 | 1.53 | 1.61 |
| Manitoba | 2025 | 11 | 7 | 235 | 1 | 1–1 | 494 | 8.5 | 1.48 | 1.54 |
| Manitoba | 2025 | 11 | 8 | 235 | 1 | 1–1 | 473 | 8.6 | 1.52 | 1.60 |
| Manitoba | 2025 | 11 | 9 | 235 | 1 | 1–1 | 455 | 8.6 | 1.56 | 1.67 |
| Manitoba | 2026 | 9 | 7 | 224 | 6 | 5–7 | 1109 | 5.8 | 1.69 | 1.61 |
| Manitoba | 2026 | 9 | 8 | 224 | 6 | 5–7 | 1068 | 5.9 | 1.75 | 1.68 |
| Manitoba | 2026 | 9 | 9 | 224 | 6 | 5–8 | 1033 | 6.0 | 1.81 | 1.74 |
| Manitoba | 2026 | 10 | 7 | 224 | 6 | 5–7 | 1075 | 5.9 | 1.74 | 1.65 |
| Manitoba | 2026 | 10 | 8 | 224 | 6 | 5–7 | 1038 | 6.0 | 1.80 | 1.72 |
| Manitoba | 2026 | 10 | 9 | 224 | 6 | 5–8 | 1007 | 6.1 | 1.86 | 1.80 |
| Manitoba | 2026 | 11 | 7 | 224 | 6 | 5–7 | 1045 | 5.9 | 1.79 | 1.70 |
| Manitoba | 2026 | 11 | 8 | 224 | 6 | 5–7 | 1013 | 6.0 | 1.85 | 1.77 |
| Manitoba | 2026 | 11 | 9 | 224 | 6 | 5–8 | 985 | 6.1 | 1.91 | 1.83 |

**Table S2 Sensitivity of provincial fit estimates to latent and infectious periods.** Minimum-BIC estimates for the Ontario 2025, Alberta 2025–2026, and Manitoba 2025–2026 waves across nine combinations of mean latent and infectious periods. Relative standard errors (RSEs) correspond to the selected fit. Accepted $I_0$ ranges span evaluated values satisfying $\Delta\text{BIC} \leq 2$ relative to the minimum within each wave and period combination; these are model-support ranges, not confidence intervals.

### 4 Latin Hypercube Sampling (LHS) paired with Partial Rank Correlation Coefficient (PRCC)

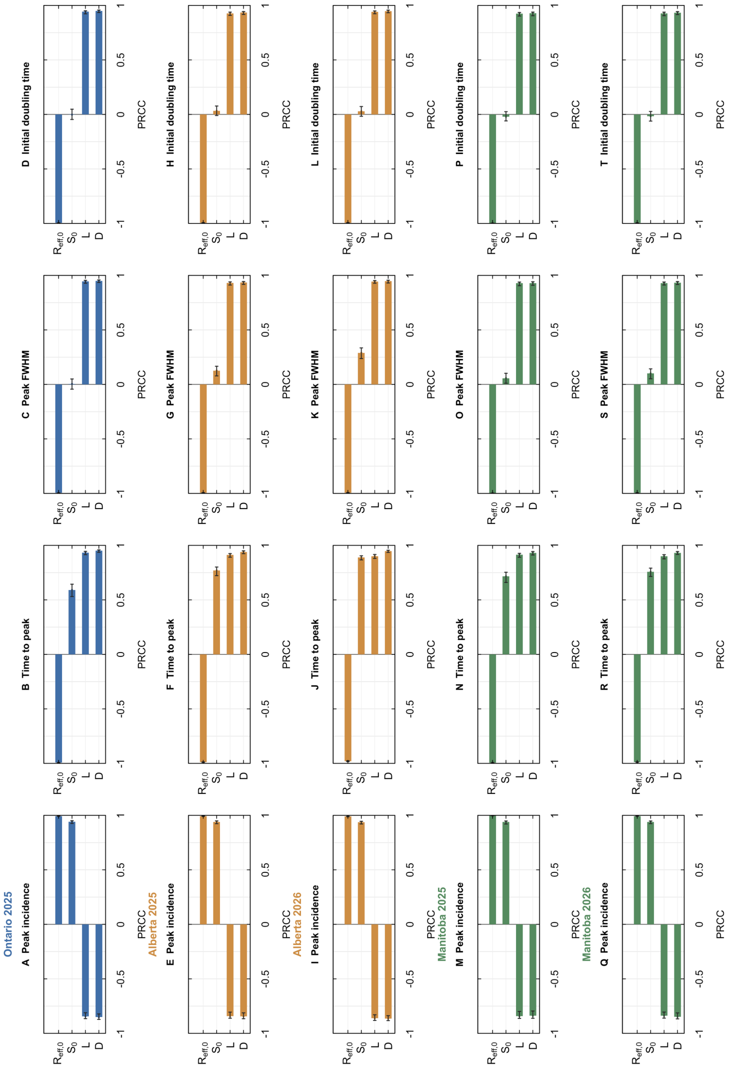


**Fig. S3 Sensitivity of provincial measles wave dynamics.** PRCCs from LHS sampling for Ontario 2025, Alberta 2025–2026, and Manitoba 2025–2026. Columns show peak incidence, time to peak, peak full width at half maximum (FWHM), and initial doubling time. Inputs were sampled within ±10% of fitted $R_{\mathrm{eff},0}$ and $S_0$, with latent periods $L = 9$–$11$ days and infectious periods $D = 7$–$9$ days; $I_0$ was held fixed. Positive and negative coefficients indicate increasing and decreasing associations, respectively, after adjustment for the other sampled inputs. Whiskers show approximate 95% bootstrap intervals.

### 5 LHS-PRCC input/output diagnostics

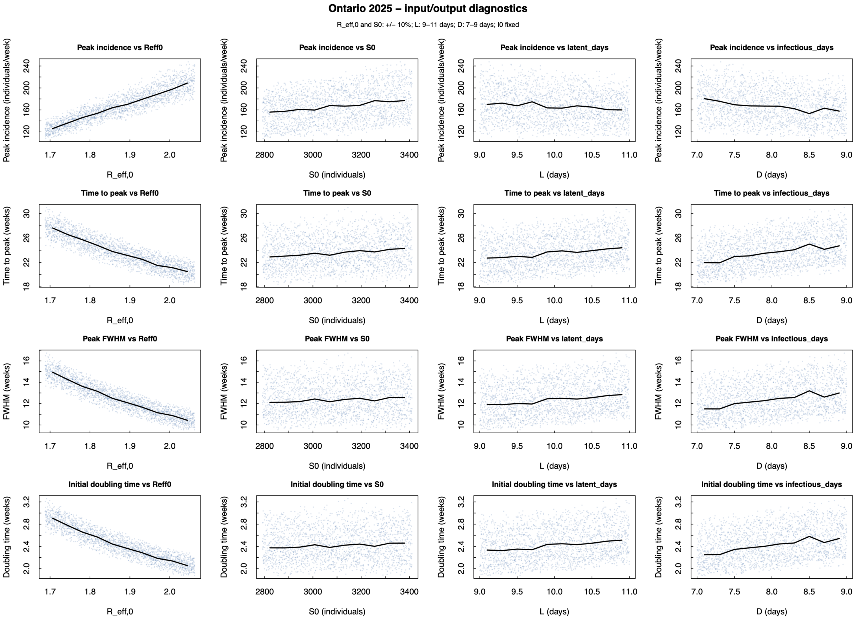


**Fig. S4 Ontario 2025 PRCC sensitivity diagnostics.** Relationships between sampled model inputs and outbreak outputs. Points represent Latin hypercube samples; black lines show median outputs within input bins. All four inputs varied simultaneously, with $I_0$ fixed.

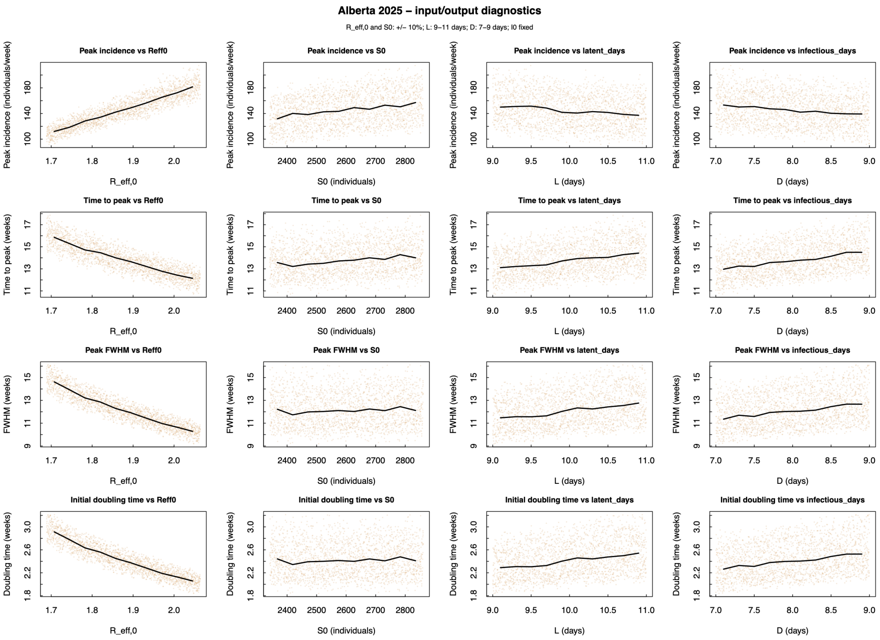


**Fig. S5 Alberta 2025 PRCC sensitivity diagnostics.** Relationships between sampled model inputs and outbreak outputs. Points represent Latin hypercube samples; black lines show median outputs within input bins. All four inputs varied simultaneously, with $I_0$ fixed.

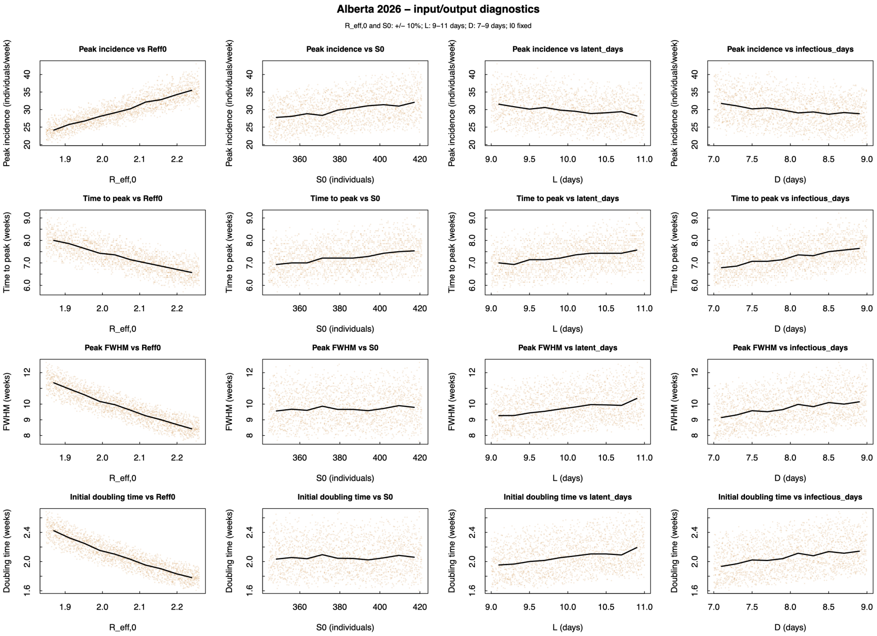


**Fig. S6 Alberta 2026 PRCC sensitivity diagnostics.** Relationships between sampled model inputs and outbreak outputs. Points represent Latin hypercube samples; black lines show median outputs within input bins. All four inputs varied simultaneously, with $I_0$ fixed.

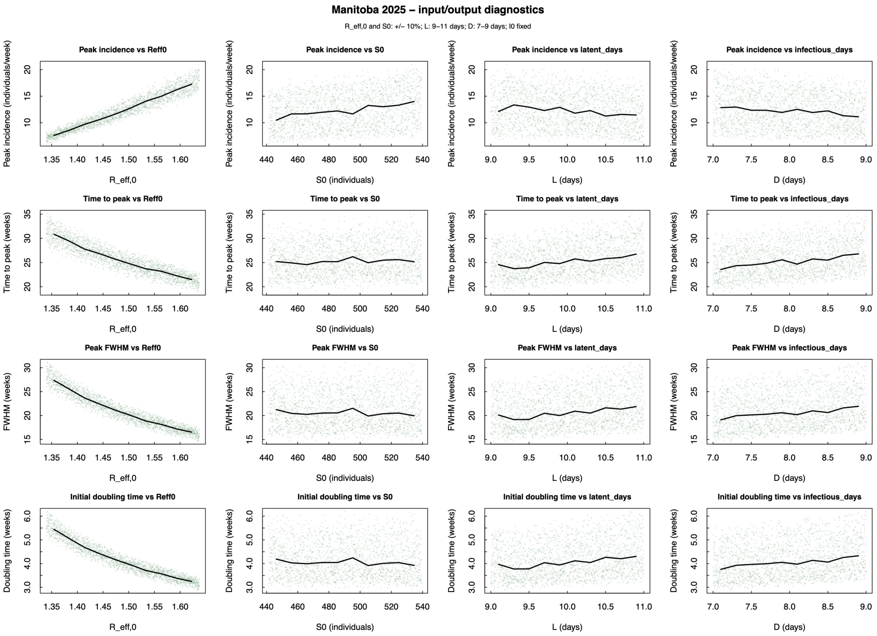


**Fig. S7 Manitoba 2025 PRCC sensitivity diagnostics.** Relationships between sampled model inputs and outbreak outputs. Points represent Latin hypercube samples; black lines show median outputs within input bins. All four inputs varied simultaneously, with $I_0$ fixed.

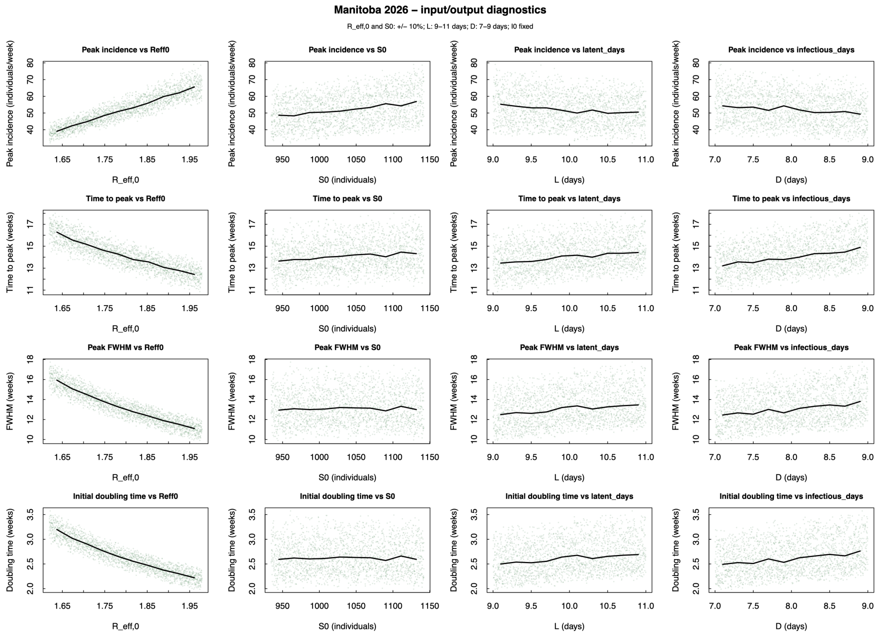


**Fig. S8 Manitoba 2026 PRCC sensitivity diagnostics.** Relationships between sampled model inputs and outbreak outputs. Points represent Latin hypercube samples; black lines show median outputs within input bins. All four inputs varied simultaneously, with $I_0$ fixed.

#### 6 Correspondance

The requests for materials should be addressed to C.S.K.
